# Death review caused by Covid 19 in Bangladesh

**DOI:** 10.1101/2022.01.23.22269626

**Authors:** Rajat Sanker Roy Biswas, Jishu Deb Nath, Fatema Emrose Nisha

**Affiliations:** Department of Medicine, Agrabad, Chittagong, CMOSHMC; Department of Medicine, CMOSHMC, Agrabad, Chittagong; CMOSHMC, Agrabad Chittagong, Bangladesh

**Keywords:** COVID-19, Co-morbidities, Outcome, Age, Hospital stay, Locality, Sex

## Abstract

**Introduction:** COVID-19 pandemic had taken away lots of human life prematurely worldwide and death laid its icy hands also on Bangladesh. So, objectives of this study were to explore the monthly distributions, age, sex, co-morbidities, localities and duration of hospital stay among the COVID death cases.

**Methods:** In this observational study six months hospital death files were collected and explored for monthly distributions, age, sex, co-morbidities, localities and hospital stay. RT-PCR positive confirmed 113 COVID deaths were enrolled and suspected COVID deaths were excluded. Ethical clearance from the hospital authority was taken before hand. Data was compiled and analyzed by SPSS-20.

**Results:** There was a low frequency of death in May-2021 and October-2021(7.1% and 2.7% respectively) but more during June -2021 to September 2021 (12.4%, 16.8%, 42.5% and 18.6% respectively). Female deaths were little more than male deaths(53.1% vs 46.9%). Age more than 51 years were the most vulnerable where 26(23%) deaths were at age group 51-60 years, 39(34.5%) deaths were at 61-70 years and 22(19.4%) deaths were more than 71 years. Mean age of death was found 60.66 years and mean duration of hospital stay was found 9.45 days. Maximum duration of hospital stay was 45 days for one patient. Co-morbidities of death cases revealed 52(46.00%) patients had DM and HTN both, 17(15.0%) patients had HTN, 16(14.1%) had DM, 3(2.6%) had BA and COPD, 4(3.5%) had CKD, 2(1.7%) had cancer, 3(2.6%) had CVD, 19(16.8%) had IHD and 16(14.1%) patients had no co-morbidities. Locality of the death cases revealed 44(38.9%) came from rural areas and 69(61.1%) came from urban areas.

**Conclusion:** Higher age group and multiple co-morbidities specially DM, HTN and IHD were related with COVID deaths mostly found in our study.

## Introduction

It is December 2021, the two years that the human race is fighting a large pandemic since 12 December 2019 caused by Severe Acute Respiratory Syndrome Coronavirus-2 (SARS CoV 2) infection termed by World Health Organization (WHO) as Coronavirus Disease, COVID-19^1^.

The WHO declared Covid-19 a global pandemic on 11March 2020. Illness ranges in severity from asymptomatic or mild to severe; a significant proportion of patients with clinically evident infection develop severe disease. Human-to-human transmission via droplets as well as through contact with fomites act as routes of the virus spread. Among the infected populations 80% are either asymptomatic or have mild disease, people have been going to their workplaces and even traveling internationally. Nevertheless, even though the virus is causing mild disease in many, the course of illness may be severe, leading to hospitalization and even death in elderly or those with comorbid conditions.^2^

Guan *et al*^2^ published a report on 1099 patients with laboratory confirmed COVID-19 from 552 hospitals in China through January 29, 2020. The most common symptoms reported were fever (43.8% on admission, and 88.7% during hospitalization) and cough (67.8%), diarrhea (3.8%) were uncommon. A severe form of the disease was reported in elderly and in patients with comorbidities. Mortality rate among diagnosed cases (case fatality rate) has a variable range; true overall mortality rate is uncertain, as the total number of cases (including undiagnosed persons with milder illness) is unknown.

In a study done in Bangladesh ^3^revealed regarding outcome of the COVID patients admitted, 85 (92.4%) patients improved, 6 (6.5%) died who were RT-PCR positive and 107 (91.15%) improved, 9 (7.7%) died who were probable cases. The death rate was little higher in our study then a study done before in Bangladesh which was 4.7%.^4^ Age and gender^5^, duration of hospital stay and localities^6^, comorbidities^7^ play some important role in the overall outcome of the Covid 19. But there are scarcity of data among the patients who died of covid 19 in the context of Bangladesh

Data of COVID are underway from the different parts of world but it is still scarce from Bangladesh regarding death revuew. Hence, objectives of this paper are to describe the Age, gender, comorbidites among the death cases of COVID 19 in a tertiary care COVID dedicated hospital of Bangladesh.

## Methods

Present study was a retrospective observational study done during a six months study period in the year 2021. In the covid unit of the hospital all death files are kept and necessary informations were collected from those retrospectively. hospital death files were explored for monthly distributions, age, sex, co-morbidities, localities and hospital stay of the deceased patients. Only RT-PCR positive confirmed cases were included in the study and a total of 113 COVID deaths were found during the study periods. Suspected COVID deaths were excluded from the study. Ethical clearance from the hospital authority was taken before hand. After compilation data was compiled and analyzed by SPSS-20.

## Results

Table 1 showing there was low frequency of death in May and October(7.1% and 2.7%) but more during June to September 2021(12.4%, 16.8%, 42.5% and 18.6%)

**Table 1:** Monthly distribution of death

| Months of year 2021 | Frequency | Percent |
| --- | --- | --- |
| May | 8 | 7.1 |
| June | 14 | 12.4 |
| July | 19 | 16.8 |
| August | 48 | 42.5 |
| September | 21 | 18.6 |
| October | 3 | 2.7 |
| Total | 113 | 100.0 |

Table 2 showing age and gender distributions of death cases where female death was more than male death(53.1% vs 46.9%). Age more than 51 years were the most vulnerable where 26(23%) were at age group 51-60 years, 39(34.5%) were at 61-70 years and 22(19.4%) were more than 71 years.

**Table 2:** Relation of sex and age group of death cases

| Age groups |  | Sex |  | Total |
| --- | --- | --- | --- | --- |
|  |  | Female | Male |  |
| Age Group | <40 years | 10(8.8%) | 3(2.7%) | 13(11.5%) |
|  | 41-50 years | 8(7.1%) | 5(4.4%) | 13(11.5%) |
|  | 51-60 years | 17(15%) | 9(8%) | 26(23%) |
|  | 61-70 years | 14(12.4%) | 25(22.1%) | 39(34.5%) |
|  | >71 years | 11(9.7%) | 11(9.7%) | 22(9.4%) |
| Total |  | 60(53.1%) | 53(46.9%) | 113(100%) |

Table 3 showing nean age was found 60.66 years and mean duration of hospital stay was found 9.45 days. Maximum duration of hospital stay was 45 days for one patient.

**Table 3:** Mean age and duration of hospital status

|  | N | Minimum | Maximum | Mean | Std.<br>Deviation |
| --- | --- | --- | --- | --- | --- |
| Age | 113 | 11 | 90 | 60.66 | 14.531 |
| Hospital stay | 113 | 1 | 45 | 9.45 | 6.981 |

Table 4 showing co morbidities of death cases where 52(46.00%) patients had DM and HTN both, 17(15.0%) patients had HTN, 16(14.1%) had DM, 3(2.6%) had BA and COPD, 4(3.5%) had CKD, 2(1.7%) had cancer, 3(2.6%) had CVD and 19(16.8%) had IHD and 16(14.1%) patients had no co morbidities.

**Table 4:**
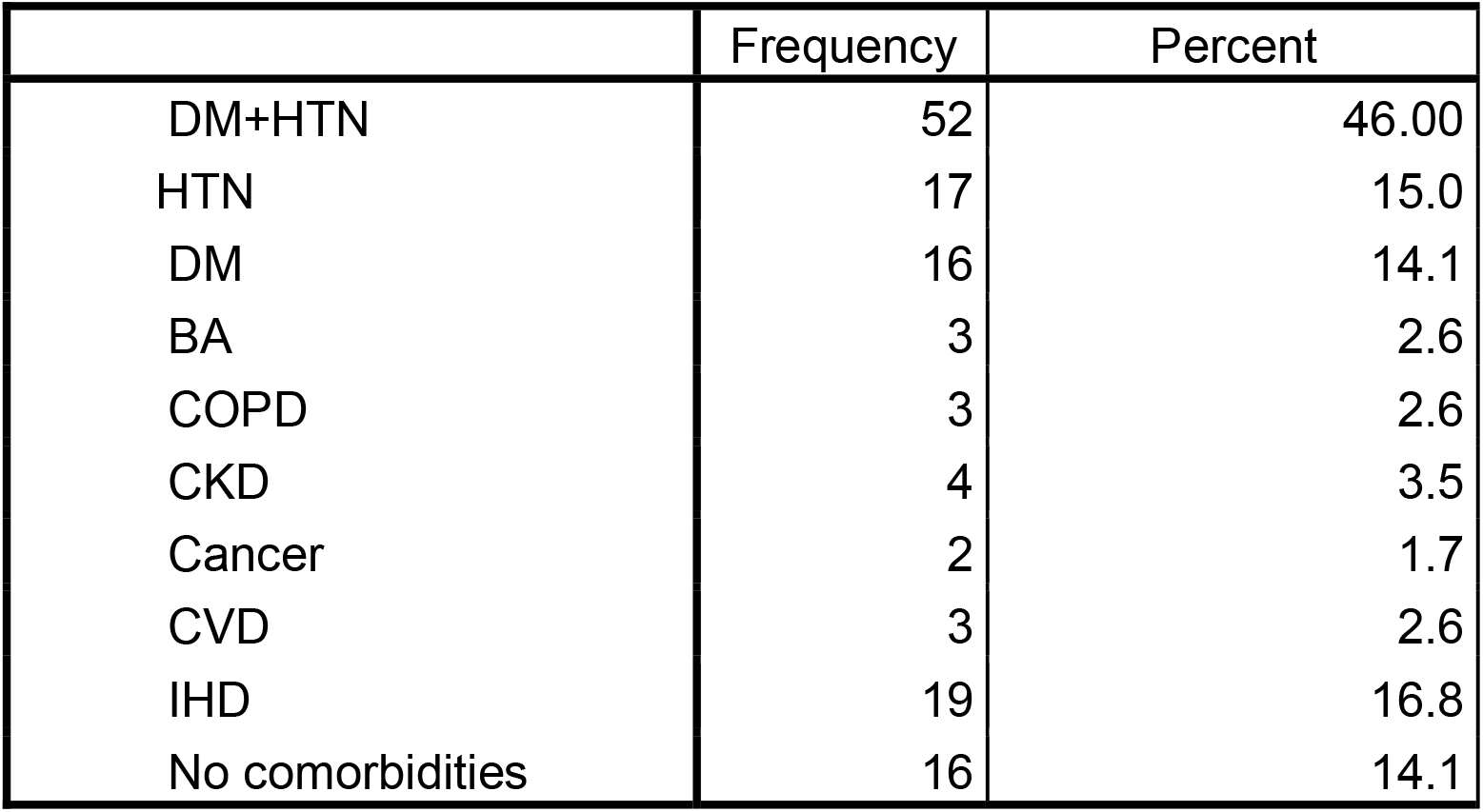
Co morbidities of the patients

|  | Frequency | Percent |
| --- | --- | --- |
| DM+HTN | 52 | 46.00 |
| HTN | 17 | 15.0 |
| DM | 16 | 14.1 |
| BA | 3 | 2.6 |
| COPD | 3 | 2.6 |
| CKD | 4 | 3.5 |
| Cancer | 2 | 1.7 |
| CVD | 3 | 2.6 |
| IHD | 19 | 16.8 |
| No comorbidities | 16 | 14.1 |

Table 5 showing locality of the death cases where 44(38.9%) came from rural areas and 69(61.1%) came from urban areas.

**Table 5:** Locality of the death cases

|  | Frequency | Percent |
| --- | --- | --- |
| Rural | 44 | 38.9 |
| Urban | 69 | 61.1 |
| Total | 113 | 100.0 |

## Discussion

There was a low frequency of death in May-2021 and October-2021(7.1% and 2.7% respectively) but more during June -2021 to September 2021 (12.4%, 16.8%, 42.5% and 18.6% respectively). Death toll was high during that time in our country and worldwide also. Second weve of Covid 19 was prevailing that time and as percentages of infection was more and frequency of death was also high.

Female deaths were little more than male deaths(53.1% vs 46.9%). But earlier study done by Huang et al^8^ and Chen et al^9^ which show 73.0% male predominance but higher than that reported by Wang et al^10^ (54.3%). This male predominance may have happened due to increased foreign travel by males for occupational or educational purposes. But in the present study death toll of female patients were little higher than the male victims and the cause is yet to be explored.

Age more than 51 years were the most Mean age of death was found 60.66 years and mean duration of hospital stay was found 9.45 days. Our findings, matched that of Asia, for example, China^11^ (median age: 47 years; 41.9% female), India^12^ (mean age 40.3 years, 66.7% male) and other reports from Bangladesh^13^ (43% were in the age range of 21–40 years, female: male ratio 1:2.33). However, studies from America^11^ (median age, 63 years) and Europe^12^ (Median age, 67.5 years) showed higher age of patients but the same male preponderance.

Co-morbidities of death cases revealed 52(46.00%) patients had DM and HTN both, 17(15.0%) patients had HTN, 16(14.1%) had DM, 3(2.6%) had BA and COPD, 4(3.5%) had CKD, 2(1.7%) had cancer, 3(2.6%) had CVD, 19(16.8%) had IHD and 16(14.1%) patients had no co-morbidities. Older adults and people of any age who have underlying medical conditions, such as hypertension and diabetes, have shown worse rognosis^14^. Diabetic patients have increased morbidity and mortality rates and have been linked to more hospitalization and intensive care unit (ICU) admissions^14,15^. People with chronic obstructive pulmonary disease (COPD) or any respiratory illnesses are also at higher risk for severe illness from COVID-19.^16^ Risk of contracting COVID-19 in patients with COPD is found to be fourfold higher than patients without COPD. There are significant differences between Bangladesh, China and the US in population demographics, smoking rates, and prevalence of comorbidities.^17^

Consistent with recent reports^8^, the percentage of patients with co morbid renal disease and malignancy was relatively low as also found in our study. Our findings have therefore added to the existing literature on the spectrum of co morbidities in patients with COVID-19 based on the larger sample sizes and representativeness of the whole patient population in Bangladesh.

Locality of the death cases revealed 44(38.9%) came from rural areas and 69(61.1%) came from urban areas. In one American study, in the rural counties, the mean prevalence of COVID-19 increased from 3.6 per 100 000 population to 43.6 per 100 000 within 3 weeks from April 3 to April 22, 2020. In the urban counties, the median prevalence of COVID-19 increased from 10.1 per 100 000 population to 107.6 per 100 000 within the same period ^18^.

Patients with co morbidities should take all necessary precautions to avoid getting infected with SARSCoV-2, as they usually have the worst prognosis. !ere is a need for a global public health campaign to raise awareness, on reducing the burden of these co morbidity illnesses causing deaths in COVID-19-infected patients.

## Data Availability

All data produced in the present study are available upon reasonable request to the authors.

## Declarations

Ethics approval and consent to participate- Taken

Consent for publication: Taken

Availability of data and material: Yes

Competing interests: None

Funding: None

Authors’ contributions: RSRB-Concept, Design, Procedure, Manuscript writing, JDN-Manuscript editing and writing, FEN-Data collection and editing

## Acknowledgements

CMOSHMC Authority

## Notes

**Financial support:** CMOSHMC

### Competing Interest Statement

The authors have declared no competing interest.

### Funding Statement

No funding

### Author Declarations

Prior permission from the Chattogram Maa Shishu O General Hospital authority was taken to conduct the study and ethical approval was also taken from the Ethical Review Board(ERB) of the same hospital.

